# From asymptomatic carriers to CADASIL: context-dependent phenotypic variability of the *NOTCH3* p.Arg1231Cys variant across three European cohorts and potential implications of LDL cholesterol

**DOI:** 10.64898/2026.10.03.26364647

**Authors:** T Nutile, D Ruggiero, M Pluntz, J Lebenberg, L Lambert, L Cipriano, A Di Pietro, V Sandroni, P Cennamo, D Hervé, S Guey, MD Calabrese, S Lucibello, AF Herzig, L Trojano, V Pizza, E Tournier-Lasserve, H Perdry, H Chabriat, AL Leutenegger, M Ciullo

**Author notes:** These authors contributed equally to the work (T Nutile, D Ruggiero, M Pluntz). These authors jointly directed this work (AL Leutenegger, M Ciullo). Corresponding authors; email: anne-louise.

## Abstract

**Background:** CADASIL caused by cysteine-altering NOTCH3 variants, the most frequent hereditary cerebral small vessel disease worldwide, is characterized by marked phenotypic variability. Mutation location within the EGFr domains of the NOTCH3 receptor has emerged as a key determinant of this variability. We investigated phenotypic variability and potential modifiers of CADASIL expression by comparing individuals harbouring the same pathogenic NOTCH3 variant across distinct recruitment settings in three European countries.

**Methods:** We compared clinical features, MRI markers, and cardiovascular risk factors among individuals harbouring the p.Arg1231Cys NOTCH3 mutation across three distinct recruitment settings: a multigenerational founder population from Cilento, Italy (CILCAD), a population-based cohort from the UK Biobank (UKB), and a clinically referred cohort from a national referral centre in Lariboisière Hospital, Paris, France (LRB). In addition, CILCAD carriers were compared with NON-Carriers from the same population. The NOTCH3 locus was finally analysed to define the shared haplotype in CILCAD and explore potential genetic contributions to phenotypic variability.

**Findings:** By comparing populations from three European countries, this study showed marked context-dependent phenotypic variability associated with the sole pathogenic p.Arg1231Cys NOTCH3 variant. In CILCAD, a multigenerational founder population, the haplotype carrying the *NOTCH3* mutation also predisposes to decreased LDL levels, resulting in an unusually low LDL cholesterol profile in this sample. Strikingly, these very low LDL levels were associated with a higher likelihood of stroke.

**Interpretation:** In European carriers of NOTCH3 p.Arg1231Cys, factors beyond mutation location, including genetic background, may substantially shape disease expression, while exceptionally low LDL levels could paradoxically be associated with greater disease severity.

**RESEARCH IN CONTEXT:** *Evidence before this study:* CADASIL, the most frequent hereditary cerebral small vessel disease worldwide, is caused by cysteine-altering *NOTCH3* mutations and is characterised by marked phenotypic variability. A position effect has emerged as a key determinant: mutations in proximal EGFr domains 1–6 associate with severe, early-onset disease, while distal domain mutations (7–34) typically result in milder phenotypes. Among distal mutations, p.Arg1231Cys is the most frequent in European populations. Beyond mutation location, hypertension and smoking have been identified as phenotypic modifiers, whereas the role of LDL cholesterol remains unclear, largely due to methodological limitations and confounding by statin use.

*Added value of this study:* This is the first study to assess phenotypic variability of a single CADASIL-causing mutation across three distinct European recruitment settings. We show that ascertainment context profoundly shapes the observed phenotype, revealing a clinical spectrum from entirely asymptomatic individuals to severe cerebrovascular disease. Genetic characterisation of the Italian founder population CILCAD identified an extended haplotype harbouring LDL-lowering alleles, establishing it as a unique model for studying low LDL in CADASIL. Strikingly, very low LDL levels were associated with a higher likelihood of stroke rather than cerebrovascular protection.

*Implications of all the available evidence:* Phenotypic variability of the p.Arg1231Cys *NOTCH3* variant extends well beyond what mutation location predicts, with ascertainment context and additional genetic or environmental modifiers playing a substantial role. These findings have direct implications for disease burden estimation, genetic counselling, and epidemiological study design. The potential deleterious association between very low LDL and stroke challenges the extrapolation of lipid-lowering targets from atherosclerotic disease to CADASIL, and warrants prospective investigation in larger longitudinal cohorts.

## Introduction

Cerebral Autosomal Dominant Arteriopathy with Subcortical Infarcts and Leukoencephalopathy (CADASIL) is the most frequent hereditary cerebral small-vessel disease caused by heterozygous cysteine-altering variants in the NOTCH3 gene^1^. These mutations typically result in an odd number of cysteine residues within one of the epidermal growth factor-like repeat (EGFr) domains of the NOTCH3 receptor^2^. The disease results from the accumulation of the extracellular domains of this protein in the wall of small vessels^3^. CADASIL is characterized by a broad spectrum of clinical manifestations, including migraine with aura, subcortical ischemic stroke, mood disorders, and cognitive decline, from isolated cognitive speed slowing up to severe dementia^4^.

Recent studies indicate that the location of cysteine-altering NOTCH3 variants within EGFr domains plays a key role in CADASIL severity, with EGFr 1–6 variants associated with the most severe and classical phenotype compared with EGFr 7–34 variants^5^. EGFr-based risk stratification has also been proposed to refine estimates of disease progression and clinical risk^6^. Nevertheless, variant location alone does not fully explain the broad clinical variability observed among patients carrying cysteine-altering NOTCH3 variants, as illustrated by the marked phenotypic heterogeneity repeatedly observed even within the same affected families. Consequently, unidentified genetic and environmental modifiers might also be involved.

The p.Arg1231Cys mutation, located in EGFr domain 31 of the NOTCH3 receptor, has been repeatedly reported to cause CADASIL in multiple small families^2,7^. Interestingly, this mutation was also found to be highly prevalent in the public database gnomAD v4.1.1 (European non-Finnish 0.38/1,000), suggesting a related exceptionally broad clinical spectrum associated with this variant, with a substantial proportion of asymptomatic or pauci-symptomatic individuals. The same variant was also found to be associated with an increased risk of incident stroke and/or transient ischemic attack (TIA)^8^ in the general population of the UK Biobank (UKB). Finally, this mutation was also found enriched in Pakistanis in association with stroke and more specifically with subcortical haemorrhagic stroke^9^.

To further investigate the phenotypic variability associated with this pathogenic p.Arg1231Cys variant and explore potential modifiers, we compared clinical, neuroimaging features and vascular risk factors across three distinct recruitment settings in Europe: 1) a multigenerational founder population from Italy (CILCAD), 2) a clinically driven cohort from a national referral centre for CADASIL in Paris, France (LRB), and 3) a population-based cohort from the UK Biobank (UKB). In the CILCAD population, complementary investigations were performed, including comparison with local non-carrier controls and analysis of a large haplotype surrounding the NOTCH3 locus.

## Subjects and Methods

The analysis was based on clinical, imaging, biological, and genetic data collected in 3 different settings

### Study samples

#### A multigenerational founder population with p.Arg1231Cys variant from Cilento (Italy) (CILCAD) and its comparative group of non-carriers from the same region

The Cardile population is one of the three isolated Italian populations (Campora, Gioi, and Cardile) included in the Cilento study^10^. Data from this population were initially collected at baseline in 2005-2008. Demographic, clinical and imaging data were additionally collected from 2020 to 2022.

Genetic data, including genotyping with microsatellite markers (deCode map) were obtained for 474 individuals and with Illumina 370K or OmniExpress SNP arrays for 569 individuals. In the whole-exome sequencing analysis of individuals from the Cilento populations, the NOTCH3 p.Arg1231Cys mutation was detected at an unexpectedly high frequency in Cardile^10^. Eighty-three p.Arg1231Cys carriers (3 homozygous and 80 heterozygous individuals) were identified by exome sequencing and/or direct genotyping of the mutation (rs201680145) in this population. Fifty-eight carriers had genotype data for microsatellite markers, and 79 (including 3 homozygous) were genotyped with SNP arrays.

In the present study, analyses were conducted in two groups: (1) the CILCAD group, comprising 83 individuals carrying the p.Arg1231Cys mutation, and (2) a comparison group of 175 NON-Carriers, adult volunteers representing the remainder of the population.

The CILCAD project was approved by the University of Naples “Federico II” Ethics Committee (reference number 191/19), and all participants provided their informed consent for participating in the study.

#### A population of patients referred to the French National Referral Centre for Cadasil in Paris at Lariboisière Hospital (LRB), France

Among the 357 index patients identified within the Lariboisière cohort of 513 individuals diagnosed with CADASIL and followed at the French National Referral Center for Rare Cerebrovascular Diseases (CERVCO, https://www.cervco.fr), 17 individuals carrying the p.Arg1231Cys mutation were recruited between 2003 and the end of 2023.

Both demographic, clinical, and imaging data were systematically collected at the time of inclusion. The study was approved by an independent ethics committee (INSERM CEEI-IRB 00003888) under the reference 17/388.

#### A population of individuals harbouring the p.Arg1231Cys variant mutation selected in the UK Biobank (UKB)

Data were obtained from the UK Biobank (UKB), a large-scale population-based prospective cohort established in 2006^11^, recruited across the United Kingdom between 2006 and 2010. After exclusion of participants who withdrew consent, 501,941 individuals, aged 40-69 years, remained available for analysis. Among them, 1,006 carried cysteine-altering NOTCH3 mutations, including 264 carriers of the p.Arg1231Cys variant. This study was conducted using the UK Biobank Resource under Application Number 94113.

### Clinical data

In patients included in the CILCAD or LRB populations, the following clinical data were systematically collected at the time of inclusion: 1) presence and age at onset of transient ischemic attacks (TIAs) or completed stroke; 2) history of migraine with or without aura; 3) global cognitive performance using the Mini-Mental State Examination (MMSE) and Mattis Dementia Rating Scale (MDRS); and 4) depressive symptoms using the Montgomery–Åsberg Depression Rating Scale (MADRS). Moreover, cognitive impairment was also considered when cognitive decline was reported by the participant or informant and supported by cognitive test scores ≥1.5 standard deviations below the reported norms, even in the absence of significant impairment in activities of daily living^12^. Dementia was diagnosed according to DSM-5 criteria^13^. Use of antidepressant or antipsychotic medication and psychotherapy were also recorded. Functional status was assessed using the modified Rankin Scale (mRS).

Smoking, alcohol consumption, hypertension (prior diagnosis, blood pressure ≥140/90 mmHg, or antihypertensive treatment), diabetes (prior diagnosis, HbA1c >6.5%, or antidiabetic treatment), and hypercholesterolemia (prior diagnosis, total cholesterol >240 mg/dL, LDL >160 mg/dL, or lipid-lowering treatment) were also systematically evaluated^14^. Finally, the total and LDL cholesterol levels were measured using the standard reference methods routinely employed by the local laboratory in both cohorts. In the CILCAD population, serum NfL concentrations were also measured using the Ella platform (Bio-Techne).

In individuals from Cardile who did not carry the p.Arg1231Cys variant, clinical information was collected using a similar but more concise approach, including at minimum a neurological questionnaire, personal and family history, clinical examination, MMSE assessment, and evaluation of cardiovascular risk factors and biological parameters using the same definitions as in the CILCAD group.

In the UK Biobank population, available data included history of stroke and dementia derived from hospital diagnosis records, primary care data, and self-report at inclusion. Vascular risk factors (hypertension, diabetes, and hypercholesterolemia) were defined using the same criteria as in the other two populations, based on medical records, self-report, physical measures at inclusion, and biological data. LDL cholesterol levels measured at inclusion were available for 254 p.Arg1231Cys carriers and 962 NOTCH3-Cys carriers. Statin use was determined from primary care prescriptions and self-report. Participants without any report of stroke or dementia from any source were considered free of these conditions.

### Imaging data

In CILCAD , cerebral MRIs were carried out using a 1.5-T system at San Luca Hospital in Vallo della Lucania. For patients referred to Lariboisière Hospital (LRB), MRI data were obtained on a 1.5T GE or 3T SIEMENS scanners. All patients underwent MRI acquisitions using 3D T1-weighted (T1w), Fluid-attenuated inversion recovery (FLAIR), and T2* MRI (or SWI MRI at 3T) sequences in both sites. The MRI sequences corresponding to data used for analysis are detailed in the Supplementary section.

In 47,415 UK Biobank participants, MRI scans were acquired at three dedicated imaging centres using identical 3T Siemens scanners (see also the Supplementary section). Imaging at time of the study (2023), MRI data were available in 99 NOTCH3-Cys carriers, 25 of them harbouring the p.Arg1231Cys variant.

Cerebral MRI lesions were assessed in 40 CILCAD participants, 17 LRB patients and 25 UK Biobank participants, all of them carrying p.Arg1231Cys. In all groups, the assessment was performed by the same expert rater (HC) using the CADA-MRIT inventory^15^. In detail, a scoring of white matter hyperintensities (WMH) of presumed vascular origin was built based on the Fazekas scale, with some modifications for the periventricular WM (score 3 was modified and score 4 was added) to take into account the very extensive areas of white matter lesions observed in CADASIL patients. The level of the deep WM did not differ from that of the Fazekas scale. The presence of WMH was assigned if a non-zero score in the periventricular, deep, or superficial WM was reported. The number of lacunes and that of microbleeds were assessed on T1 and on T2*/SWI sequences, respectively. The presence of brain atrophy was defined as a non-zero score for deep or superficial atrophy. The presence or absence of WMH, of at least one lacune, one microbleed, or brain atrophy was considered separately^15^.

### Data analysis

#### Comparison between the different populations harbouring the p.Arg1231Cys variant

Comparisons between the CILCAD group, the LRB group, and the UK Biobank (UKB) populations were performed using linear or logistic regression models, depending on the outcome. Age (at inclusion or MRI, according to the timing of the variable assessed) and sex were included as covariates, except when age or sex itself was the variable of interest. MMSE analyses were further adjusted for education level. Vascular risk factors (hypertension, hypercholesterolemia) were added for a sensitivity analysis when comparing clinical and imaging outcomes. Each model included the CILCAD sample and the relevant comparison population (LRB or UKB), with group membership entered as a binary variable; statistical significance was assessed using Wald tests associated with this variable. In rare cases of model degeneration due to complete separation (i.e., all or no individuals presenting the condition in one group, such as “at least one vascular risk factor” in LRB or dementia in UKB), score tests were used instead. Associations between LDL cholesterol levels and stroke history were examined using logistic regression models adjusted for prespecified covariates. All analyses excluded missing data.

#### Complementary analysis in the CILCAD population

Age and sex differences were first assessed directly, whereas analyses of biochemical parameters, modifiable cardiovascular risk factors, MMSE scores, stroke history, and cognitive impairment were adjusted for age and sex. Comparisons of genetic data between the CILCAD group and NON-Carriers from Cardile were performed using linear mixed models (LMMs) accounting for familial relationships, with the *lmekin* function of the *coxme* R package (version 2.2.22, https://CRAN.R-project.org/package=coxme).

### Estimation of the age of the ancestor

The age of the most recent common ancestor of the p.Arg1231Cys carriers was estimated in the population using the Estiage program^16^ under a stepwise model at the different markers. In detail, the Estiage estimates the age of the mutation using a set of haplotypes of genetic markers, the reconstructed ancestral haplotype, and the allelic frequencies of the marker alleles. The age estimation was carried out using 58 CILCAD and microsatellite marker data on chromosome 19 as required by program implementation at the time of the analysis. The microsatellite D19S179 was chosen as the target marker because it is closest to the rs201680145 (p.Arg1231Cys) variant. Haplotypes were reconstructed moving left and right from the target marker, and for each marker, the ancestral allele was determined as being the most frequent allele among carriers. In the presence of ties, the ancestral allele could not be determined, and the haplotype reconstruction was not carried out further. Haplotype reconstruction was possible for 9 markers upstream of the target (D19S179) and 8 markers downstream, spanning 67.7 cM, with D19S179 approximately in the middle of the haplotype. For each marker, the allele frequency was estimated from 606 NON-Carriers from the same population. The Estiage program provides maximum likelihood estimates of the number of generations since the most recent common ancestor using multi-locus marker data information on carriers. It assumes that all carriers in the sample descended from a common ancestor who introduced p.Arg1231Cys n generations ago. An estimate of n is obtained from the size of the haplotype shared by individuals on both sides of the disease locus by finding the most likely positions of recombination on the ancestral haplotype in the different carriers; then, the value of n is converted to age in years by multiplying by the assumed 25 years per generation. The chronological date was determined using the year 1955, the mean of the birth dates of all carriers was included in the analysis.

### Haplotype shared between patients

Maternal and paternal chromosomes in 569 Cardile samples, including 79 CILCAD (76 heterozygous and 3 homozygous individuals), were reconstructed by phasing 3,523 SNPs on chromosome 19 from Illumina SNP array (call rate >=95% and MAF >= 1%) and using a linkage disequilibrium (LD)-based phasing algorithm, implemented in the SHAPEIT2 program (v2.r904) with the “duohmm” option that harnesses parent-offspring or duo information for phasing^17^. The 82 mutation-carrying chromosomes were used to estimate the ancestral haplotype on the short arm of chromosome 19 by assigning to each SNP the most frequent allele in those chromosomes. The length of the haplotype shared between mutation-carrying chromosomes was defined as the region around the NOTCH3 mutation, which includes all those SNPs carrying the ancestral allele in the analyzed chromosomes. In detail, in mutation-carrying chromosomes, for each SNP, the number of chromosomes with an allele different from that in the ancestral haplotype was counted so that zero corresponds to positions where all the 82 mutation-carrying chromosomes share the ancestral allele, 1 to positions where only one mutation-carrying chromosome differs, etc. For comparison purposes, the same procedure was also applied to the 76 alternative chromosomes not carrying the mutation. In each of the two groups, consisting of the 82 mutation-carrying and the 76 alternative-allele-carrying chromosomes from the 79 CILCAD, allele differences between chromosomes were counted at each SNP. Allele differences detected between mutation-carrying chromosomes were markedly reduced in a large region around the NOTCH3 gene, as compared to the alternative chromosomes (Figure 1).

**Figure 1:**
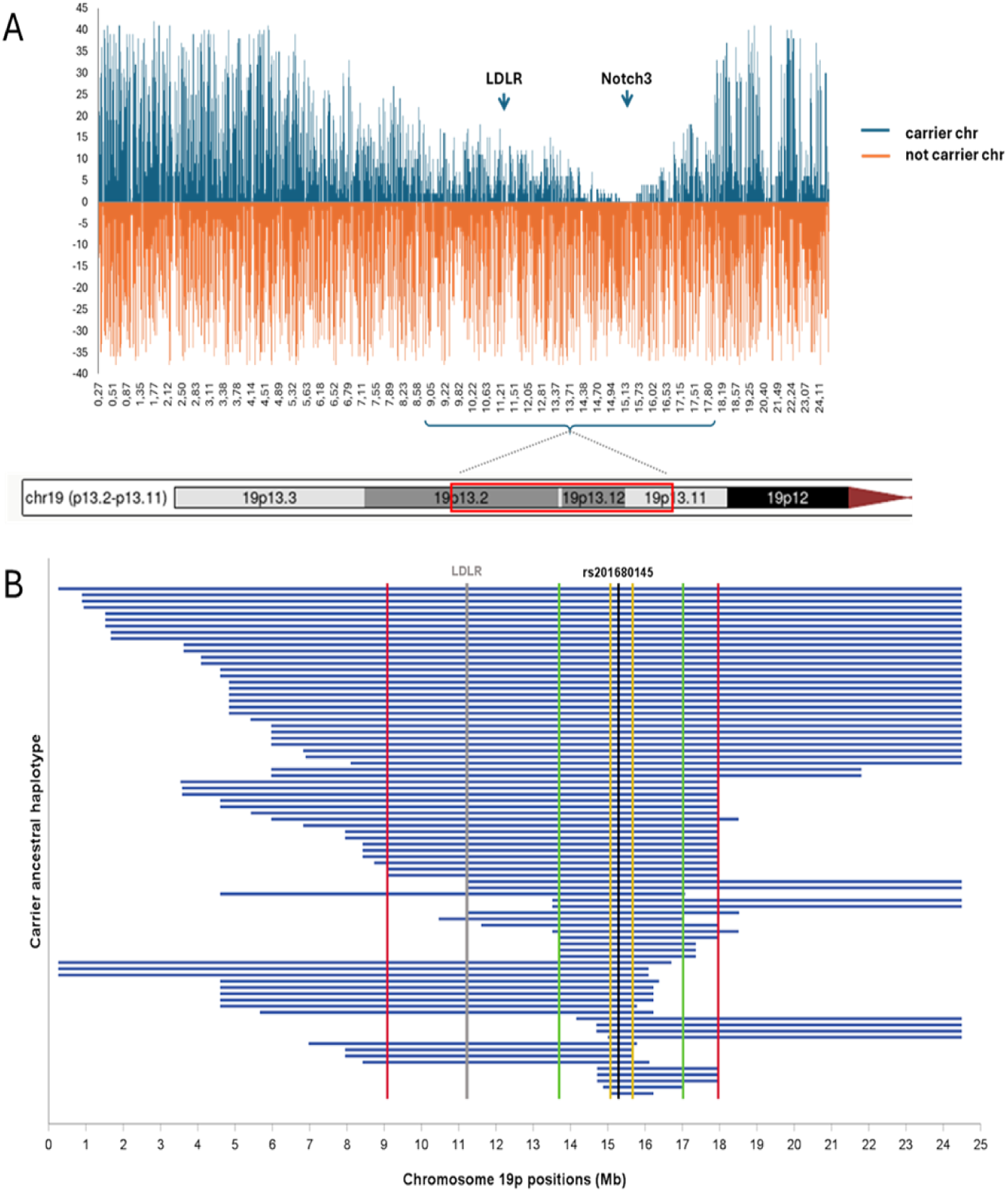
Haplotype sharing. **A.** Graphical representation of the short arm of chromosome 19 in mutation-carrying chromosomes (in blue) and alternative allele-carrying chromosomes (in orange) of CILCAD participants. The X-axis represents SNP positions; the Y-axis reports the number of chromosomes carrying an allele different from the ancestral allele (for the mutation-carrying chromosomes) or from the more frequent allele (for the alternative chromosomes). The shape of the upper part of the graph highlights that the length of the shared haplotype varies according to the number of mutation-carrying chromosomes. The region around the NOTCH3 gene is shared by all mutation-carrying chromosomes. **B.** Haplotype sharing among the 82 mutation-carrying chromosomes. The black line indicates the position of the p.Arg1231Cys mutation (rs201680145 variant), the red lines delimitate the 8.9 Mb haplotype shared by the 57.3% of chromosomes, the green lines delimitate the 3.3 Mb haplotype shared by the 73.1% of chromosomes, and the yellow lines delimitate the 600 Kb haplotype shared by all chromosomes. The grey line indicates the position of the LDLR gene. Haplotypes are ordered starting from the top to the bottom: those sharing the 8.9 Mb region (red lines), those sharing th 3.3 Mb (green lines), and those only sharing the 600 Kb region (yellow lines).

### Polygenic score for LDL levels

A polygenic score (PGS) for LDL was computed for 79 CILCAD and 490 NON-Carriers from the Cardile population, and for 23 LRB. We explored the PGS Catalog and selected the LDL PGS and metadata reported by Zhang et al.^18^ (PGS003784), as it was the most recent LDL PGS for European population ancestry. Thus, genotypes of the 3,490 genome-wide SNPs reported by Zhang H et al. (GW-LDL-PGS) or the 77 SNPs located in the shared haplotype region (8.9Mb-LDL-PGS) (Supplementary Figure 1) were used to estimate PGS. Differences in PGS were assessed by a 2-sided independent samples t-test. Similarly, a haplotype-LDL-PGS based on the presence of the effect allele was estimated for each of the two individual chromosomes. Haplotype-PGS difference was calculated between mutation-carrying chromosomes and alternative-allele-carrying chromosomes.

## Results

### Differences between the three populations with the p.Arg1231Cys variant

Although CILCAD subjects carried the same NOTCH3 mutation and shared necessarily a large homogeneous genetic and environmental background, they exhibited large phenotypic variability. Out of the 83 carriers, 52 individuals already presented with at least one of the classical clinical manifestations of CADASIL (symptomatic individuals). Thirteen of these symptomatic individuals were under 50 years of age. Sixteen individuals had suffered from attacks of migraine with or without aura, 16 had presented a stroke; 40 suffered from mood disturbances, mainly depression; finally, 8 presented with cognitive impairment at the time of their clinical assessment, 4 of whom were demented. Eight individuals were asymptomatic, and 5 of them were under 50 years. This variability remained obvious even in individuals aged over 50 years.

Imaging data were available in 40 participants (32 symptomatic and 8 asymptomatic). Of those, 26 had a non-zero WMH score in deep, periventricular, or superficial WM; 12 presented at least with one lacune; two had at least one microbleed. Brain atrophy was detected in 13 participants (Table 1, Supplementary Figure 2). Among individuals aged over 50 years, 47.8% patients had lacunes, and 91.7% WMHs.

**Table1:** Population statistics of p.Arg1231Cys carriers in the CILCAD, LRB, and UKB samples and p-values for their comparisons.

|  | Population samples |  |  | Comparison (p-value) |  |
| --- | --- | --- | --- | --- | --- |
|  | CILCAD [N = 53] | LRB [N = 17] | UKB [N = 264] | CILCAD - LRB | CILCAD - UKB |
| Age, mean ± SD | 56.0 ± 20.4 [53] | 62.1 ± 9.5 [17] | 55 ± 8.1 [264] | p = 0.244 <sup>(1)</sup> | p = 0.539 <sup>(1)</sup> |
| Age at imaging, mean ± SD | 54.0 ± 20.5 [40] | 62.1 ± 9.5 [17] | 63.1 ± 7.5 [25] | p = 0.127 <sup>(1)</sup> | **p = 0.037** <sup>(1)</sup> |
| Female sex, n (%) | 29 (54.7 %) [53] | 9 (52.9 %) [17] | 138 (52.3 %) [264] | p = 0.898 <sup>(1)</sup> | p = 0.745 <sup>(1)</sup> |
| Education level in years, median (Q1-Q3) | 9 (5 - 13) [48] | 12 (9 - 15.5) [16] | 11 (10 - 13) [197] | **p = 0.002** | p = 0.103 |
| Hypertension, n (%) | 21 (53.8 %) [39] | 16 (94.1 %) [17] | 166 (62.9 %) [264] | p = 0.053 | p = 0.458 |
| Hypercholesterolemia, n (%) | 12 (22.6 %) [53] | 11 (64.7 %) [17] | 132 (51.6 %) [256] | **p = 0.004** | **p = 4x10<sup>-5</sup>** |
| Total Cholesterol (mg/dL), mean ± SD | 176.9 ± 38.6 [53] | 169.6 ± 38 [16] | 215.8 ± 42.6 [254] | p = 0.451 | **p = 2x10<sup>-9</sup>** |
| - In subjects taking statins, mean ± SD | 172.1 ± 28.3 [9] | 152.4 ± 36.7 [8] | 179.5 ± 39 [53] | p = 0.266 | p = 0.689 |
| - In subjects not taking statins, mean ± SD | 177.9 ± 40.6 [44] | 186.8 ± 32.7 [8] | 225.4 ± 38.2 [201] | p = 0.767 | **p = 1x10<sup>-12</sup>** |
| LDL (mg/dL), mean ± SD | 99.9 ± 31.7 [53] | 101.7 ± 29.6 [16] | 136.48 ± 32 [254] | p = 0.963 | **p = 4x10<sup>-13</sup>** |
| - In subjects taking statins, mean ± SD | 97.11 ± 20.8 [9] | 88.94 ± 32.7 [8] | 109.3 ± 28.9 [53] | p = 0.509 | p = 0.446 |
| - In subjects not taking statins, mean ± SD | 100.5 ± 33.6 [44] | 114.46 ± 21 [8] | 143.6 ± 28.8 [201] | p = 0.354 | **p = 2x10<sup>-16</sup>** |
| Statin use, n (%) | 9 (17 %) [53] | 9 (52.9 %) [17] | 55 (20.8 %) [264] | **p = 0.007** | **p = 0.023** |
| Diabetes, n (%) | 9 (17 %) [53] | 3 (17.6 %) [17] | 38 (16.4 %) [232] | p = 0.963 | p = 0.414 |
| Smoking, n (%) | 14 (32.6 %) [43] | 7 (41.2 %) [17] | 125 (47.5 %) [263] | p = 0.443 | p = 0.063 |
| Alcohol, n (%) | 23 (53.5 %) [43] | 9 (52.9 %) [17] | 34 (12.9 %) [263] | p = 0.735 | **p = 1x10<sup>-8</sup>** |
| At least 1 risk factor, n (%) | 37 (84.1 %) [44] | 17 (100 %) [17] | 237 (90.8 %) [261] | p = 0.182 <sup>(2)</sup> | p = 0.738 |
| At least 1 stroke, n (%) | 7 (13.2 %) [53] | 12 (70.6 %) [17] | 5 (1.9 %) [264] | **p = 1x10<sup>-4</sup>** | p = 0.751 |
| MMSE, mean ± SD | 29 (27 - 30) [49] | 25 (23 - 29) [17] | NA | **p = 0.003** <sup>(3)</sup> | NA |
| mRS > 1, n (%) | 1 (2.6 %) [38] | 13 (76.5 %) [17] | NA | **p = 0.002** | NA |
| MMSE ≤ 24, n (%) | 7 (14.3 %) [49] | 6 (35.3 %) [17] | NA | **p = 0.015** <sup>(3)</sup> | NA |
| Dementia | 2 (3.8 %) [53] | 2 (11.8 %) [17] | 0 (0 %) [264] | p = 0.155 | p = 0.196 <sup>(2)</sup> |
| **CADAMRIT data** |  |  |  |  |  |
| Total WMH score, median (Q1-Q3) | 1.5 (0 - 3) [40] | 9 (8 - 10) [17] | 2 (1.75 - 4) [24] | **p = 6x10<sup>-16</sup>** | p = 0.469 |
| At least one lacune, n (%) | 12 (30.8 %) [39] | 14 (82.4 %) [17] | 8 (32 %) [25] | **p = 0.004** | p = 0.63 |
| At least one microbleed, n (%) | 2 (5.6 %) [36] | 12 (70.6 %) [17] | 5 (20.8 %) [24] | **p = 0.001** | p = 0.068 |
| Deep or superficial atrophy, n (%) | 13 (32.5 %) [40] | 15 (88.2 %) [17] | 12 (48 %) [25] | **p = 0.002** | p = 0.494 |

For each variable, the number of individuals with available data is reported in square brackets. P-values correspond to the Wald test associated with each variable, adjusting for age (at imaging for CADA-MRIT variables and at inclusion otherwise) and sex, with the following exceptions: (1) not adjusted for age or sex; (2) score test used because the model required for the Wald test did not converge; (3) additionally adjusted for education level. P-values < 0.05 are shown in bold

Despite this large variability also detected in the 3 cohorts, individuals selected from population-based samples (CILCAD, UKB) exhibited a lower clinical severity than patients recruited from a referral centre (Table 1). Specifically, stroke prevalence was significantly higher in LRB (70.6%) than in CILCAD (13.2%); meanwhile, CILCAD exhibited a higher prevalence than UKB (1.9%), though this difference did not reach statistical significance. Accordingly, CADA-MRIT parameters (including WMH, lacunes, microbleeds, and atrophy) align with a more severe phenotype in LRB and reflect a closer clinical picture between CILCAD and UKB. These results also hold when adjusting the comparisons on hypercholesterolemia and hypertension (Supplementary Table 1)

### Complementary results obtained in the CILCAD population

A higher prevalence of stroke was observed in CILCAD compared to NON-Carriers from the same population. Furthermore, an even larger prevalence of stroke was detected in CILCAD individuals aged over 50 years (28.3% vs 7.4%, OR=4.92; 95% CI 2.31-10.51; p<0.0001). The prevalence of cognitive impairment was also higher in CILCAD, and the MMSE score was lower than in the NON-Carriers (Table 2). In this comparison, data from different sources were used for stroke and dementia: neurological examination, family history, and additional clinical data already available from previous follow-ups in the Cilento study.

**Table 2.** Neurological and cardiovascular risk factors data of CILCAD individuals and NON-Carriers from the Cardile population.

|  | CILCAD [N = 65] | NON-Carriers [N = 175] | p-value |
| --- | --- | --- | --- |
| Female sex, n (%) | 37 (56.9%) [65] | 101 (57.7%) [175] | 0.91 |
| Age, years (mean ± SD) | 60.8 ± 21.5 [65] | 55.3 ± 17.1 [175] | **0.04** |
| **<u>Cognitive Functions</u>** |  |  |  |
| MMSE (mean ± SD) | 27.6 ± 3.4 [49] | 28.9 ± 2.0 [172] | **1.2x10<sup>-5</sup>** |
| Cognitive Impairment, n (%) | 8 (13.3%) [60] | 7 (4.0%) [175] | **0.033** |
| NfL, pg/ml (mean±SD) | 21.3 ± 32.2 [50] | 13.4 ± 21.8 [175] | **0.026** |
| Stroke, n (%) | 16 (26.7%) [60] | 3 (1.7%) [175] | **4.1x10<sup>-10</sup>** |
| **<u>Cardiovascular Risk Factors</u>** |  |  |  |
| BMI (mean ± SD) | 26.7 ± 4.4 [39] | 26.9 ± 4.6 [174] | 0.62 |
| Total Cholesterol, mg/dL (mean ±SD) | 176.9 ± 38.6 [53] | 204.2 ± 40.2 [175] | **6.3x10<sup>-5</sup>** |
| LDL, mg/dL (mean ±SD) | 99.9 ± 31.7 [53] | 119.2 ± 33.4 [175] | **1.3x10<sup>-3</sup>** |
| Statin Consumption, n (%) | 9 (17.0%) [53] | 34 (19.4%) [175] | 0.64 |
| Hypercholesterolemia, n (%) | 12 (22.6%) [53] | 65 (37.1%) [175] | **0.04** |
| Glucose, mg/dL (mean ± SD) | 100.1 ± 19.5 [53] | 99.8 ± 27.0 [175] | 0.92 |
| HbA1c, % (mean ± SD) | 5.82 ± 0.63 [53] | 6.00 ± 0.77 [175] | 0.14 |
| Type 2 Diabetes, n (%) | 9 (17.0%) [53] | 25 (14.3%) [175] | 0.84 |
| Hypertension, n (%) | 21 (53.8%) [39] | 96 (60.0%) [160] | 0.58 |
| Alcohol, n (%) | 23 (53.5%) [43] | 92 (53.2%) [173] | 0.88 |
| Smoking, n (%) | 14 (32.5%) [43] | 43 (25.1%) [171] | 0.35 |

Biological analyses showed that CILCAD individuals had lower levels of total and LDL cholesterol than NON-Carriers; no other differences were detected in other vascular risk factors. Levels of NfL were significantly increased in CILCAD compared to NON-Carriers (Table 2).

### Age estimate of the ancestor carrying the p.Arg1231Cys mutation in Cardile

Haplotype reconstruction indicates that the mutation is of very recent origin, dating it approximately 6 generations ago (estimated between 1855–1885). The identification of a very long ancestral haplotype (43 Mb), with large portions of which are still found in most CILCAD, is consistent with a very recent origin of the mutation. In a previous study, we demonstrated that Cardile and Gioi, another Italian isolated population, share a common origin and became genetically distinct over generations because of the isolation following their separation^19^. Therefore, the absence of the p.Arg1231Cys mutation in Gioi is consistent with its appearance in Cardile after their separation, either via a *de novo* mutation or a founder effect from a nearby Italian individual.

### Shared CILCAD Haplotype: a segment encompassing LDL-Lowering Genetic Variants

The frequency of the p.Arg1231Cys mutation in the NOTCH3 gene was already found to be exceptionally high in the population of Cardile (14.5%)^10^. A total of 83 individuals carrying the p.Arg1231Cys mutation (CILCAD group) were identified, all belonging to a single 1,862-member pedigree spanning 17 generations (Supplementary Figure 3). A large haplotype around the NOTCH3 mutation was found to be shared by most of the mutation-carrying chromosomes. Indeed, 57.3% of the chromosomes (Nchr=47) share a haplotype of 8.9 Mb (chromosome region 9-18Mb), the shared haplotype was shortened to 3.3 Mb in 73.1% of chromosomes (Nchr=60), and to about 600 Kb in 100% of mutation-carrying chromosomes (Nchr=82). In alternative chromosomes, the count of allele differences is uniformly noisy along the entire short arm of chromosome 19 (Figure 1).

A lower polygenic score for LDL (GW-LDL-PGS) was observed in CILCAD compared with NON-Carriers (p=0.011). This difference was highly significant for 8.9Mb-LDL-PGS (p=1.6×10^-18^) (Figure 2A). According to the hypothesis by which LDL-lowering alleles were carried by the same chromosome carrying the p.Arg1231Cys mutation, a haplotype polygenic score (haplo-LDL-PGS) for each of the two individual chromosomes was calculated in CILCAD and NON-Carriers. We found that chromosomes carrying the p.Arg1231Cys allele have a lower haplo-LDL-PGS than those carrying the alternative allele from both CILCAD (p=3.94×10^-4^) and NON-Carriers (p=3.99×10^-18^), confirming the presence of LDL-lowering alleles on the same chromosome carrying the p.Arg1231Cys mutation (Figure2B). Overall, these results confirm that the haplotype carrying the p.Arg1231Cys mutation encompasses one or more LDL-lowering genetic variants.

**Figure 2:**
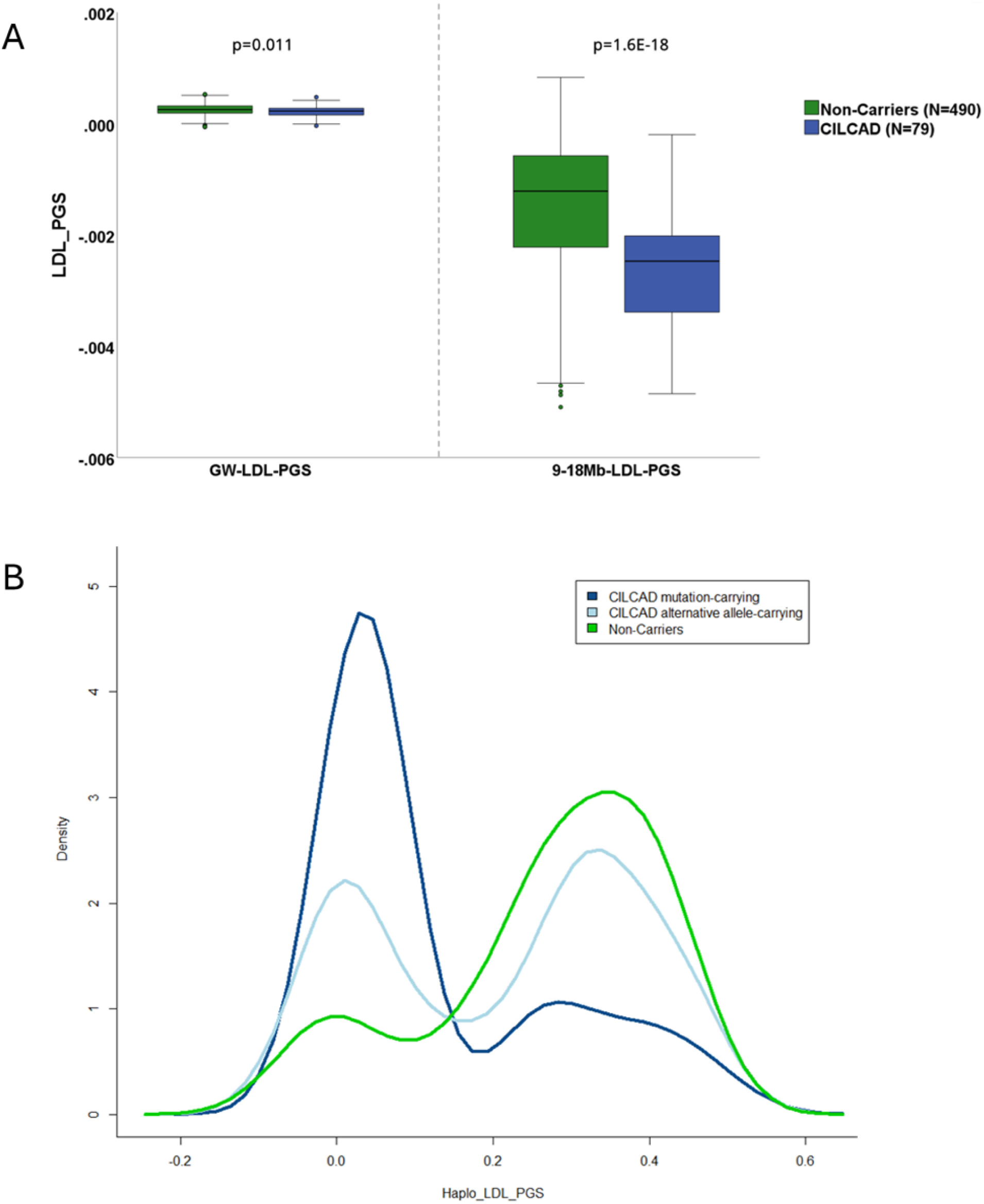
Polygenic score for LDL levels in CILCAD and NON-Carriers from the Cardile population. **A.** PGS estimated with 3,490 genome-wide SNPs and PGS estimated with 77 SNPs in the 8.9 Mb of the shared region. **B.** Density plot of cholesterol haplotype-PGS in mutation-carrying chromosomes of CILCAD (in blue), in alternative allele-carrying chromosomes of CILCAD (in light blue) and both chromosomes from NON-Carriers (in green).

### LDL CHOLESTEROL LEVELS ACROSS THE THREE POPULATIONS AND POTENTIAL IMPACT ON THE PHENOTYPE

In agreement with the results reported above between carriers and NON-Carriers in the Cardile population, hypercholesterolemia was found to be less frequent, and LDL cholesterol levels were lower in CILCAD compared to UKB or LRB populations. This difference vanished in individuals taking statin treatments (Table 1).

Accordingly, the CILCAD sample had a lower 8.9Mb-LDL-PGS than the LRB sample (p=2.6×10^-6^) (Supplementary Figure 4), whose individuals did not share the haplotype around the NOTCH3 gene. The proportion of subjects aged over 50 years and having LDL levels below 80 mg/dL (and not taking statins) was also much larger among CILCAD individuals (20% vs 0.9%; N=25; mean age=69.2 years) than in the entire UKB dataset (carriers and non-carriers; N=266,099; mean age=59.9 years),

Given the disease variability and the genetically determined lower LDL levels characterizing the CILCAD sample, we searched for a possible link between circulating LDL level and ischemic manifestations (stroke and lacunes on MRI) in this specific population. A negative association was detected between LDL levels and stroke in CILCAD, after adjustment for age, sex, and statin treatment (p=3.7×10^-3^). Stroke events were mainly observed in subjects with extremely low levels of LDL cholesterol (86% of individuals with a positive history of stroke had LDL cholesterol levels below 80 mg/dL) (Figure 3A). This association remained significant when hypertension and smoking status were considered (p=0.021). In a sensitivity analysis, this association was also statistically significant (p=0.012) in the subset of subjects who were never treated by statins. In this group, the LDL mean value was 65.7 mg/dL in patients who experienced a stroke and 103.9 mg/dL in subjects who did not (Figure 3B). To further investigate this finding, we analysed previous LDL cholesterol data obtained in 2005 in the same individuals, prior to any stroke occurrence. Stroke events occurred more frequently in individuals with lower LDL levels measured 18 years earlier (overall: p=0.036; no-statin subgroup: p=0.02), showing that the association between low LDL levels and stroke was unlikely to be explained by post-stroke lifestyle changes.

**Figure 3:**
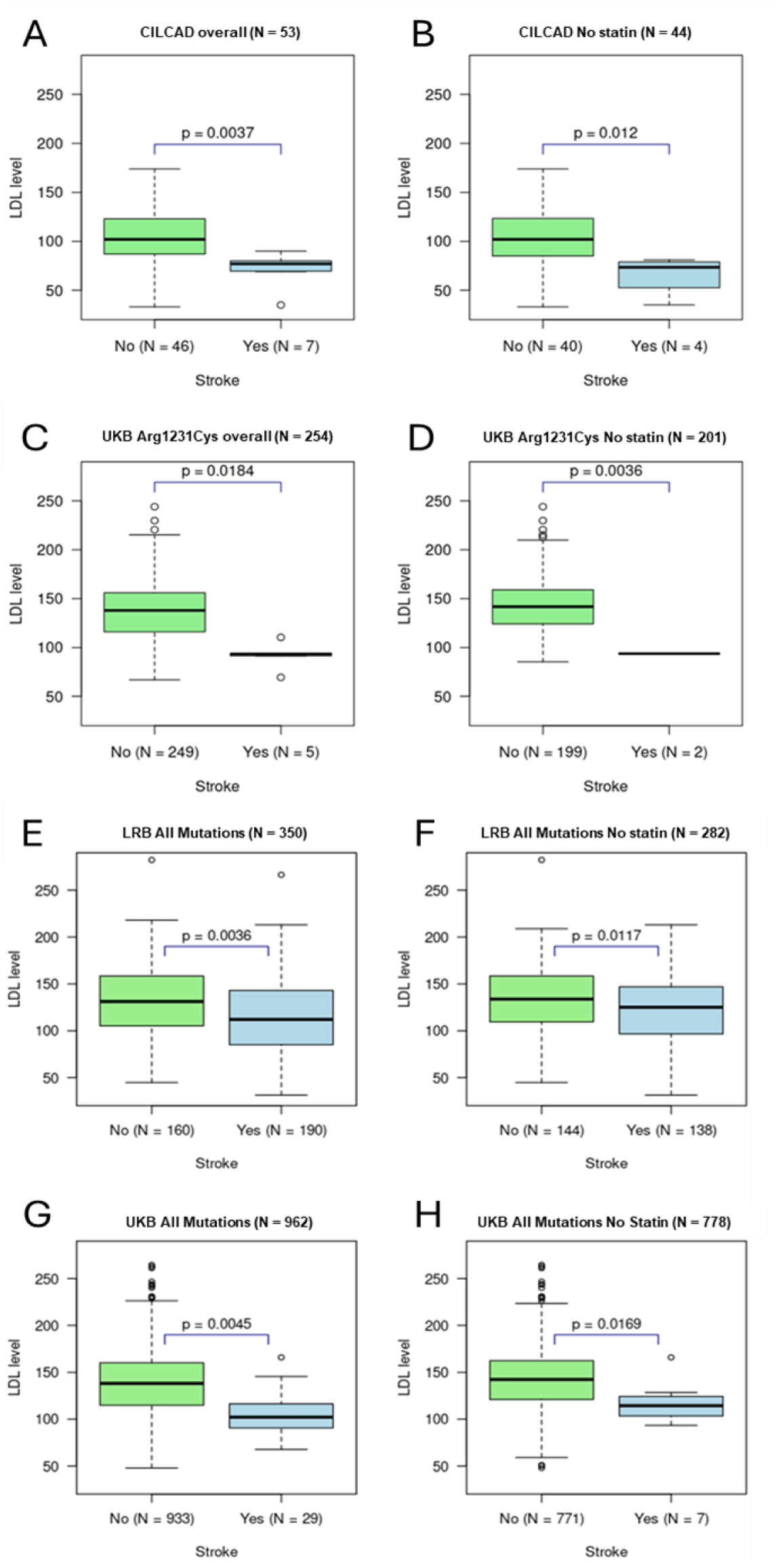
Box plots of LDL level according to prevalent stroke in overall CILCAD (A) or in the subgroup not taking statins (B), in the overall UKB p.Arg1231Cys mutation carriers (C) or in the subgroup not taking statins (D), and in all NOTCH3 mutation carriers, (overall and subgroups not taking statins), in LRB (E-F) and UKB (G-H) samples. P-values are from score tests of logistic regressions of Stroke on LDL adjusted for age, sex, and statin treatment (overall only)

In the UKB, although LDL levels were globally higher than in the CILCAD group, we observed that patients with stroke had lower LDL levels in p.Arg1231Cys carriers (p=0.018) than patients without stroke and that such a difference was even more significant in the subgroup not taking statins (p=3.6×10^-3^) (Figure 3C-D). The same results were obtained after adjusting for hypertension and smoking status (overall p=4.5×10^-3^; no-statin group p=0.013). A similar difference was found in the UKB sample including all NOTCH3 cysteine-altering mutations (N=962, p=4.5×10^-3^), in the embedded subgroup of individuals who did not take statins (N=778, p=0.017), as well as after further adjusting for hypertension and smoking status in addition to age and sex (overall p=5.3×10^-3^; no-statin group p=0.016).

In the LRB cohort, no conclusion could be drawn due to the small sample size of p.Arg1231Cys mutation carriers (overall N=16; not taking statins, N=8). In the whole group of LRB patients, including all NOTCH3 cysteine-altering mutations, LDL cholesterol was also found lower in patients with a positive history of stroke: (N=350, p=3.6×10^-3^). This difference was still significant in individuals without statins (N=282, p=0.011) (Figure 3E-H), and after adjusting for hypertension and smoking status in addition to age and sex (overall p=0.011; no-statin group p=0.019).

A negative association was found between LDL levels and the presence of lacunes in LRB overall (N=350, p=3.3×10^-3^) and in the no-statin group (N=282, p=6.0×10^-4^). Differently, no association was found in CILCAD (overall N=39, p=0.42; no-statin group N=31, p=0.94) and in the UKB (overall N=93, p=0.80; no-statin group N=79, p=0.91), probably because of a lack of power due to smaller sample sizes (Supplementary Figure 5A-B).

## Discussion

A key finding of this study is that recruitment setting fundamentally shapes the clinical phenotype expressed by carriers of the p.Arg1231Cys *NOTCH3* variant, one of the most prevalent CADASIL-causing mutations identified in the general population to date. Despite harbouring the same cysteine-altering variant, individuals exhibited substantial heterogeneity in both clinical presentation and neuroimaging manifestations, demonstrating that mutation location alone is insufficient to predict disease expression. Participants identified through population-based recruitment, as in the UK Biobank, or within the isolated Cilento founder population, displayed markedly milder phenotypes than patients referred to a national referral centre, a finding consistent with the well-recognised ascertainment bias inherent to clinic-based studies of rare genetic diseases. This three-setting comparison provides a distinctive epidemiological opportunity to disentangle ascertainment effects from true biological variability, by contrasting unselected individuals from the general population, members of a multigenerational Italian founder population, and patients diagnosed across different French regions and referred to an expert centre for rare diseases. As anticipated, those recruited through the referral pathway exhibited the most severe and classic form of CADASIL phenotype with the highest burden of clinical and neuroimaging abnormalities, while population-identified participants presented substantially milder, often pauci-symptomatic or asymptomatic profiles. The intermediate phenotype observed in the CILCAD founder cohort with a slightly higher prevalence of stroke manifestations further suggests that disease expression is shaped not only by the causal variant, but also by ascertainment context and, potentially, by additional genetic or environmental modifiers.

Another major result is that such phenotypic contrasts can also be detected in a population as the CILCAD founder population carrying the p.Arg1231Cys NOTCH3 mutation. Although the overall phenotype observed in this cohort did not differ significantly from that observed in the UK Biobank, the CILCAD population nonetheless revealed striking within-group contrast, spanning a large clinical spectrum from entirely asymptomatic individuals to those with severe manifestations such as stroke and cognitive impairment. This breadth of expression, observed among individuals sharing not only the same *NOTCH3* cysteine-altering mutation but also a common ancestral background through multigenerational transmission, reinforces the well-established but incompletely understood phenotypic heterogeneity of CADASIL^20,21^, and strongly suggests that individual-level genetic modifiers, distinct from the shared ancestral background, as well as environmental exposures or epigenetic mechanisms, are likely contributors to the clinical diversity associated with the p.Arg1231Cys *NOTCH3* variant.

In the context of persistent phenotypic variability within the CILCAD founder population, the unexpected difference in LDL cholesterol levels between mutation carriers and NON-Carriers from the Cardile population raised the question of whether LDL cholesterol may act as a modifier of disease expression in this cohort. CILCAD participants exhibited significantly lower LDL levels than carriers of the p.Arg1231Cys variant in both the hospital-based LRB cohort and the UK Biobank. Our analyses reveal that this distinctive lipid profile is embedded in the genetic architecture of the region itself. The large 8.9 Mb *NOTCH3* haplotype shared by CILCAD subjects also harbours LDL-lowering alleles (including variants at the *LDLR* locus) as captured by both a lower LDL polygenic score and a haplotype-specific LDL polygenic score. This feature was absent in the LRB cohort and the UK Biobank. This clearly establishes CILCAD as a genetically unique model for investigating the long-term impact of constitutively low LDL levels on CADASIL disease expression.

Paradoxically, we observed a negative association between LDL cholesterol and stroke in the CILCAD population. Strikingly, participants in this group who had experienced a stroke presented with mean LDL levels below 1.8 mmol/L or 70 mg/dL, values that approximate or fall below the thresholds currently recommended for patients at very high cardiovascular risk^22^. Furthermore, the analysis of previously collected LDL data in CILCAD suggests that this association was unlikely to be driven by reverse causation. The fact that such low LDL concentrations were nonetheless associated with a higher likelihood of stroke in this non-atherosclerotic small vessel disease population underscores a fundamentally different, and potentially inverse, relationship between LDL cholesterol and cerebrovascular injury in CADASIL compared with classical atherosclerotic disease, and raises important questions about the extrapolation of lipid-lowering treatment targets to this patient population.

While traditional vascular risk factors such as smoking and hypertension have been repeatedly associated with greater disease severity or earlier stroke onset in CADASIL^23,24^, the role of LDL cholesterol has remained largely unclear. Much of the uncertainty stems from a key methodological limitation: most existing studies have relied on binary definitions of hypercholesterolaemia based on lipid thresholds and medication use, rather than examining LDL as a continuous quantitative variable. Although Dupé et al. reported an association between hypercholesterolaemia, stroke occurrence, and functional dependency in CADASIL^20^, the majority of subsequent studies found no significant correlation with disease outcomes^24–34^. Interpreting these discrepant findings is further complicated by the progressive broadening of statin indications with current guidelines suggesting lipid-lowering therapy for virtually all patients with ischaemic stroke regardless of aetiology, with LDL targets set below 70 mg/dL for those at high vascular risk^22^. As a result, observed LDL levels in patient-based CADASIL cohorts increasingly reflect treatment effects rather than intrinsic biological variation, substantially confounding any attempt to disentangle the true relationship between LDL cholesterol and cerebrovascular outcomes in this disease. Interestingly, in the present study, we also observed that among cysteine-variant carrier populations of the LRB cohort and the UK Biobank, patients who experienced at least one stroke event had a cholesterol level close to 100 mg/dL but lower than in individuals with no history of ischemic stroke. These results were found after adjustment for age, sex, and the use of statins. Interestingly, all these results align with recent observations in the broader form of sporadic SVD, suggesting a “paradoxical” inverse association, where lower LDL levels correlate with worse clinical outcomes, including an increased risk of stroke^35^. Indeed, while extensive evidence supports LDL as a causal risk factor for atherosclerotic large-vessel ischemic stroke, a subset of studies focusing specifically on SVD have reported a negative or null association, where lower LDL levels are linked to a higher risk of recurrent stroke or worse SVD progression. From a broader perspective, discrepancies in the literature regarding the role of LDL in stroke may be attributed to differences in cohort ethnicity, stroke subtypes, the specific pathological context, and the confounding effect of statin use^36–39^. Obviously, longitudinal research in larger samples is essential to define the role of LDL in CADASIL as a specific SVD condition and to clarify the therapeutic implications of lipid management in these patients.

In conclusion, this study demonstrates that disease expression in CADASIL is profoundly influenced by ascertainment context and suggests that additional genetic or environmental modifiers may contribute to the striking phenotypic variability observed among carriers of the same *NOTCH3* cysteine-altering variant. We believe that these findings have direct implications for how the clinical spectrum and true population burden of hereditary cerebral small vessel diseases should be assessed and underscore the value of population-based epidemiological approaches in this field. Furthermore, the paradoxical association between very low LDL cholesterol levels and worse cerebrovascular outcomes challenges the assumption that lipid-lowering strategies proven beneficial in atherosclerotic disease can be straightforwardly extended to CADASIL and calls for a critical reassessment of lipid management in this population.

## Data Availability

The data used in this study originate from three sources. UK Biobank data are available to bona fide researchers through the UK Biobank data access procedures. Data from the Cilento population study and individual-level patient data collected at the French National Referral Center for Rare Cerebrovascular Diseases (CERVCO, AP-HP) are not publicly available due to privacy, ethical and legal restrictions. Access to these data may be considered upon reasonable request, subject to applicable regulations and approval by the relevant institutions.

## CONTRIBUTORS

ALL and MC developed the study design; ALL and MC were responsible for funding acquisition. MC, TN, and DR coordinated data collection of CILCAD participants in the municipality; VP performed clinical examinations of CILCAD participants and coordinated MRI acquisition at the San Luca Hospital; LC and ADP performed neurological examinations in the Cardile population; MDC and SL collected neuropsychometric data in the CILCAD cohort; LT analysed neuropsychometric tests in the CILCAD cohort. JL was responsible for MRI data gathering, quality control, and imaging measurements. TN, DR, and MP were involved in data cleaning, statistical analysis, data visualization, and interpretation of the results. PC and AFH contributed to the genetic data analysis. HP contributed to the methodological development of genetic data analysis. ETL supervised the clinical genetic diagnosis of CADASIL for all individuals investigated in the French cohort. DH and SG performed the clinical evaluation of all individuals investigated in the French cohort. LL and VS participated in the comparative analyses of data. HC was responsible for the coordination and clinical supervision of the French cohort. All authors had full access to the data. MC, ALL, DR, and HC wrote the original draft. All authors critically reviewed and commented on the paper, approved the final version of the manuscript, and were responsible for the decision to submit the manuscript for publication.

## ACKNOLEDGEMENTS

We express our sincere gratitude to the Cardile population for their participation in the study. We are sincerely thankful to Dr Michelina De Cristofaro for her contribution in clinical data collection in Cardile; the San Luca Hospital in Vallo della Lucania, for the support in the clinical data collection and MRI examinations for the CILCAD cohort; Dr Giovanni D’Arena for biochemical measurements; Rosita Manna and Lina Scelza for scheduling medical examination appointments in Cardile. We thank Dr Paola Italiani and Dr Daniela Melillo for their support in ELLA-based NfL measurements. We thank Dr Francesco Habetswallner for critical discussions.

We are deeply grateful to Mrs Fanny Fernandes and Abbas Taleb for their invaluable support in study coordination, regulatory procedures, data collection, and data cleaning, and to Mrs Carla Machado and Aude Jabouley for their important contribution to the neuropsychological evaluation of all French patients. We sincerely thank the CADASIL-France association for their continuous support and valuable contribution to the development and maintenance of the French CADASIL cohort. We also acknowledge the support of ARNEVA and the IHU-VHBI in Bordeaux for their continued contribution to maintaining and advancing the cohort study and its associated database.

The work was supported by the Alzheimer Association (AARG-NTF-20-683992); the Agence Nationale de la Recherche (ANR-20-CE17-0031); the “National Biodiversity Future Center” on ‘Biodiversity’, financed under the National Recovery and Resilience Plan (NRRP), Mission 4, Component 2, Investment 1.4 “Strengthening of research structures and creation of R&D ‘national champions’ on some Key Enabling Technologies”; the CNR projects FOE-2021-DBA.AD005.225 and FOE 2022 - DSB.AD006.371.

## Conflict of Interest

The Authors declare no conflicts of interest

## Funding

The project was funded by the Alzheimer’s Association (AARG-NTF-20-683992) and Agence Nationale de la recherche (ANR-20-CE17-0031)

## Notes

### Competing Interest Statement

The authors have declared no competing interest.

### Author Declarations

- Ethics committee/IRB of University of Naples Federico II gave ethical approval for this work under reference number 191/19 - Ethics committee/IRB of Institut national de la sante et de la recherche medicale (INSERM CEEI-IRB 00003888) gave approval for this work under the reference 17/388

