## Supplemental File for "From asymptomatic carriers to CADASIL: context-dependent phenotypic variability of the *NOTCH3* p.Arg1231Cys variant across three European cohorts and potential implications of LDL cholesterol"

### Supplementary data

MRI sequences used for imaging data collection are detailed below.

In both the CILCAD and LRB studies and for data acquired at 1.5T: T1w-MRI were sagittally acquired with a voxel size of 0.8x0.8 or 1.0x1.0 mm2, with TR of 1700, 1900, or 1950 or 9000 ms, TI of 0, 776, 859 or 900ms, TE of 1.9, 2.6, 6.6, 6.8 or, 6.9 or 2.6 ms, a slice thickness of 1, 1.6, 3.5, or 4 or 1mm and a space between slices of 0, 0.8, 4.2, or 4.8, 4.2 or 0 mm; FLAIR MRI were axially or sagittally acquired with a voxel size of 0.5x0.5, 0.7x0.7 or 1.0x1.0 mm2, with TR of 5000, 7990 or, 8500 or 5000 ms, TI of 210, 1600, 2370 or, 2440 or 1600ms, TE of 96, 140-160 or 340 ms, a slice thickness of 1.1, 3.5,  4 or 5.5 or 1.1mm and a space between slices of 0, 4.2 4.8 or 5.5, 4.2 or 0 mm; T2* MRI were axially acquired with a voxel size of 0.5x0.5 or 0.9x0.9 mm2, with TR of 35, 465, or 601500-600, or 35 ms, TE of 15 or 25 ms, a slice thickness of 3.5, 4, or 5 or 5.5 mm, and a space between slices of 4.82, 4.28, or 5 or 5.5 mm.

For data acquired at 3T in the LRB study: T1w-MRI were sagittally acquired with a voxel size of 0.4x0.4 or 1.0x1.0 mm2, with TR of 1800-2200 ms, TI of 900-910 ms, TE of 2.3 or 3.2 ms, a slice thickness of 0.4 or 1 mm and a space between slices of 0 mm; FLAIR MRI were sagittally acquired with a voxel size of 0.5x0.5, or 1.0x1.0 mm2, with TR of 5000-6000 ms, TI of 1600-1900 or 2200 ms, TE of 375-395 ms, a slice thickness of 0.9 or 1.1 mm and a space between slices of 0 mm; SWI MRI were sagittally acquired with a voxel size of 0.4x0.4 mm2, with TR of 22-27 ms, TE of 15 or 20 ms, a slice thickness of 1.5 or 2 mm, and a space between slices of 0 mm.

In the UKB study, the standardized protocol included T1-weighted MPRAGE (1×1×1 $mm^3$ resolution, TR=2000ms, TE=2.01ms, TI=880ms, flip angle=8°) and T2-weighted FLAIR sequences (1.05×1.0×1.0 $mm^3$ resolution, TR=5000ms, TE=395ms, TI=1800ms). The MRIs were performed, on average, 11 years after inclusion.

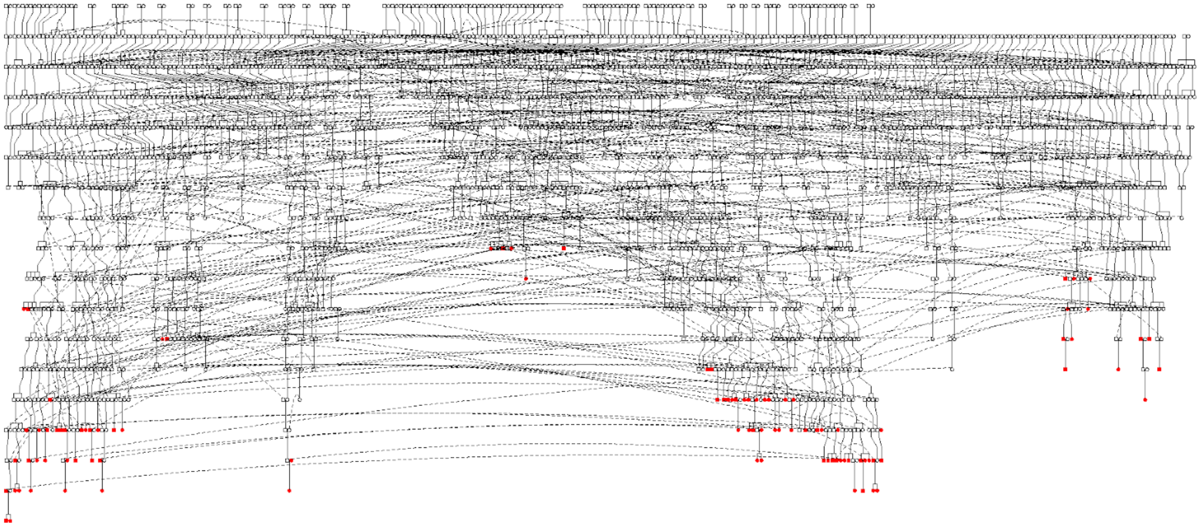

***Supplementary Figure 1.*** *17-generation pedigree connecting all individuals carrying the p.Arg1231Cys mutation in the Cardile population. Dashed lines indicate individuals duplicated for a pedigree drawing purpose. CILCAD participants carrying the p.Arg1231Cys mutation are reported in red.*

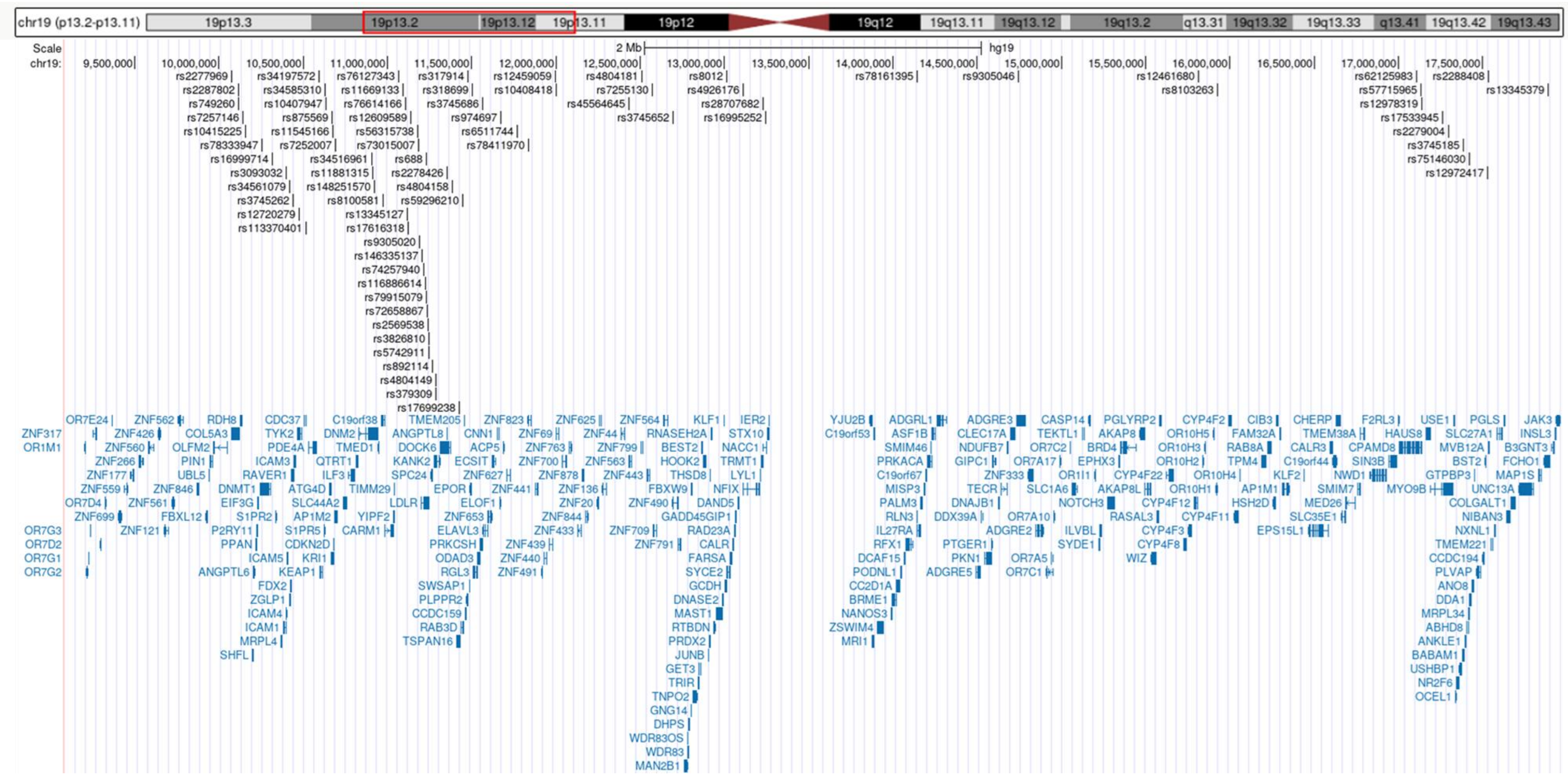


***Supplementary Figure 2.*** *Representation of the 77 SNPs in the shared haplotype region and included in the LDL-PGS-8.9Mb with respect to the genes in the region, according to the UCSC Genome Browser and GENECODE v49lift37.*

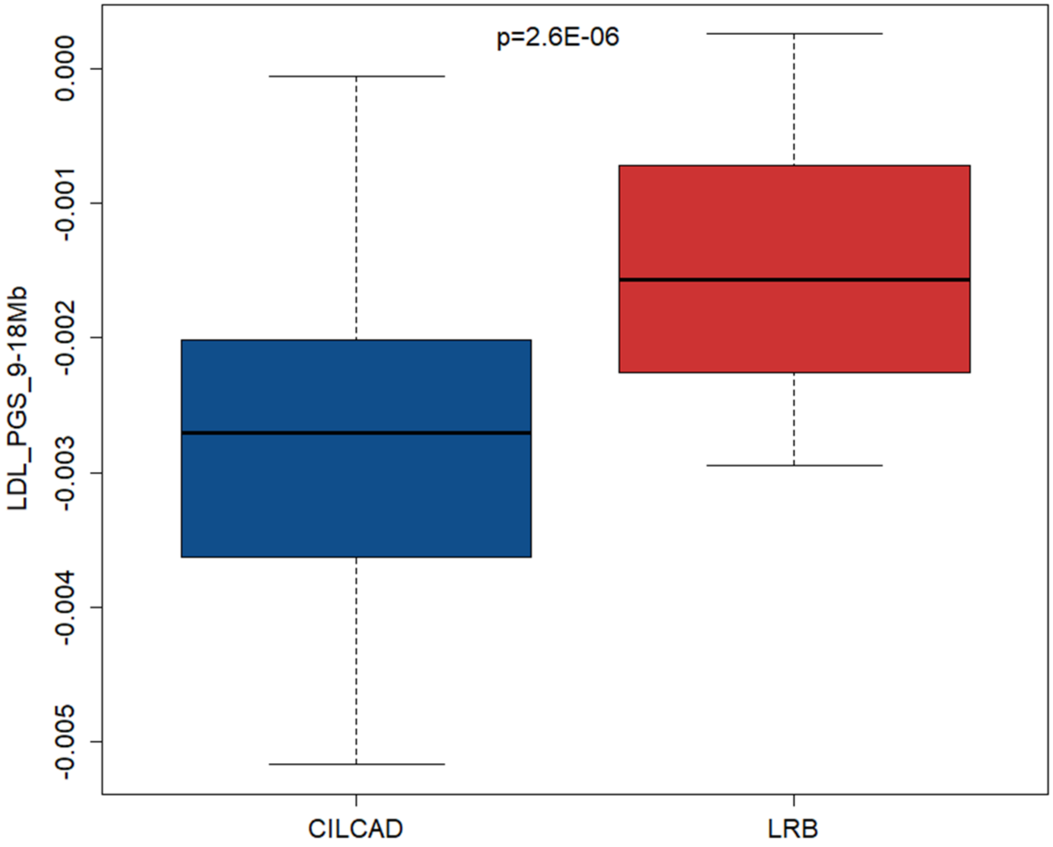


***Supplemenatry Figure 3.*** *Polygenic score for LDL levels estimated with 77 SNPs in the 8.9 Mb of the shared region in CILCAD and LRB.*

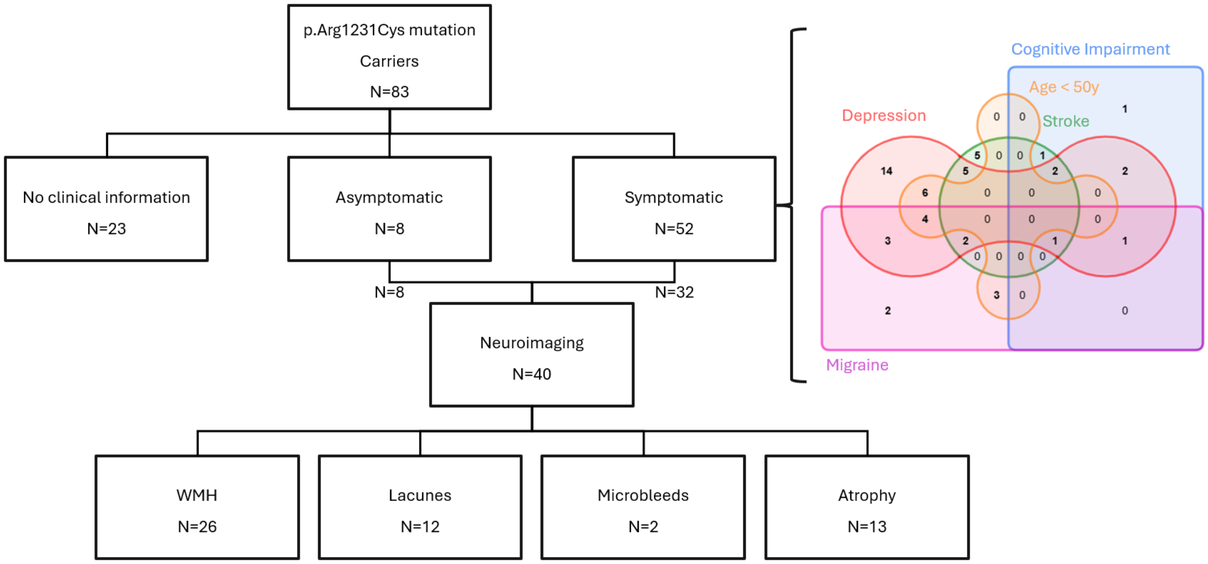


***Supplementary Figure 4.*** *Clinical and neuroimaging data of the p.Arg1231Cys mutation Carriers (CILCAD). The number of individuals with MRI features (presence of WMH, at least one lacune, at least one microbleed, presence of atrophy) was reported.*

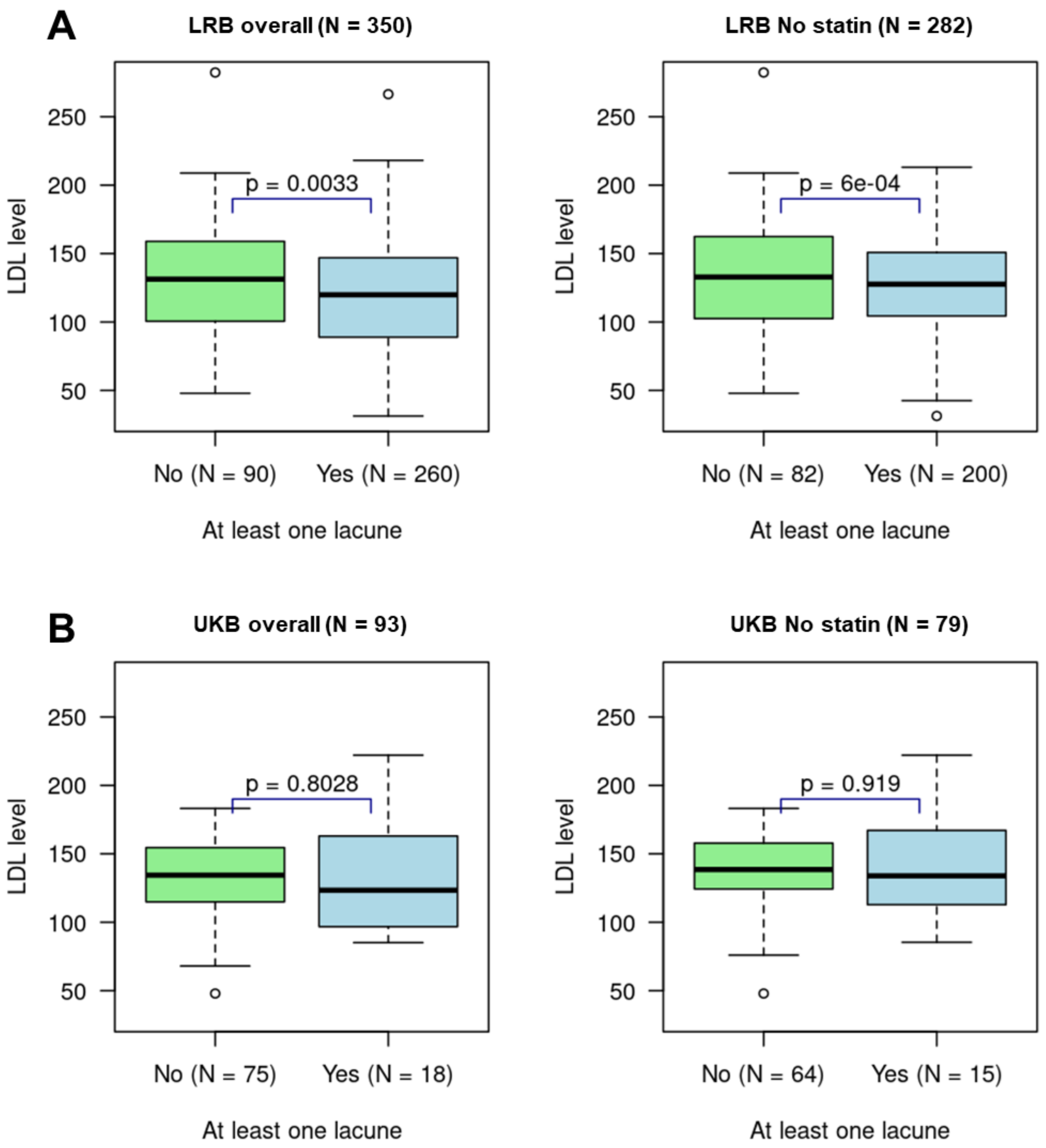


***Supplementary Figure 5.*** *Box plots of LDL levels according to the presence of lacunes in all NOTCH3Cys mutation carriers (overall and subgroups not taking statins) in LRB (A) and UKB (B) samples. Score tests, adjusted for age, sex, and statin treatment (overall only)*

***Supplementary Table 1****: Population statistics of p.Arg1231Cys carriers in the CILCAD, LRB, and UKB samples and p-values for their comparisons.*

| | Population samples | | | Comparison (p-value) | |
|---|---|---|---|---|---|
| | **CILCAD [N = 53]** | **LRB [N = 17]** | **UKB [N = 264]** | **CILCAD - LRB** | **CILCAD - UKB** |
| **Age, mean ± SD** | 56.0 ± 20.4 [53] | 62.1 ± 9.5 [17] | 55 ± 8.1 [264] | p = 0.244 (1) | p = 0.539 (1) |
| **Age at imaging, mean ± SD** | 54.0 ± 20.5 [40] | 62.1 ± 9.5 [17] | 63.1 ± 7.5 [25] | p = 0.127 (1) | **p = 0.037** (1) |
| **Female sex, n (%)** | 29 (54.7 %) [53] | 9 (52.9 %) [17] | 138 (52.3 %) [264] | p = 0.898 (1) | p = 0.745 (1) |
| **Education level in years, median (Q1-Q3)** | 9 (5 - 13) [48] | 12 (9 - 15.5) [16] | 11 (10 - 13) [197] | **p = 0.002** | p = 0.103 |
| **Hypertension, n (%)** | 21 (53.8 %) [39] | 16 (94.1 %) [17] | 166 (62.9 %) [264] | p = 0.053 | p = 0.458 |
| **Hypercholesterolemia, n (%)** | 12 (22.6 %) [53] | 11 (64.7 %) [17] | 132 (51.6 %) [256] | **p = 0.004** | **p = $4x10^{-5}$** |
| **Total Cholesterol (mg/dL), mean ± SD** | 176.9 ± 38.6 [53] | 169.6 ± 38 [16] | 215.8 ± 42.6 [254] | p = 0.451 | **p = $2x10^{-9}$** |
| **- In subjects taking statins, mean ± SD** | 172.1 ± 28.3 [9] | 152.4 ± 36.7 [8] | 179.5 ± 39 [53] | p = 0.266 | p = 0.689 |
| **- In subjects not taking statins, mean ± SD** | 177.9 ± 40.6 [44] | 186.8 ± 32.7 [8] | 225.4 ± 38.2 [201] | p = 0.767 | **p = $1x10^{-12}$** |
| **LDL (mg/dL), mean ± SD** | 99.9 ± 31.7 [53] | 101.7 ± 29.6 [16] | 136.48 ± 32 [254] | p = 0.963 | **p = $4x10^{-13}$** |
| **- In subjects taking statins, mean ± SD** | 97.11 ± 20.8 [9] | 88.94 ± 32.7 [8] | 109.3 ± 28.9 [53] | p = 0.509 | p = 0.446 |
| **- In subjects not taking statins, mean± SD** | 100.5 ± 33.6 [44] | 114.46 ± 21 [8] | 143.6 ± 28.8 [201] | p = 0.354 | **p = $2x10^{-16}$** |
| **Statin use, n (%)** | 9 (17 %) [53] | 9 (52.9 %) [17] | 55 (20.8 %) [264] | **p = 0.007** | **p = 0.023** |
| **Diabetes, n (%)** | 9 (17 %) [53] | 3 (17.6 %) [17] | 38 (16.4 %) [232] | p = 0.963 | p = 0.414 |
| **Smoking, n (%)** | 14 (32.6 %) [43] | 7 (41.2 %) [17] | 125 (47.5 %) [263] | p = 0.443 | p = 0.063 |
| **Alcohol, n (%)** | 23 (53.5 %) [43] | 9 (52.9 %) [17] | 34 (12.9 %) [263] | p = 0.735 | **p = $1x10^{-8}$** |
| **At least 1 risk factor, n (%)** | 37 (84.1 %) [44] | 17 (100 %) [17] | 237 (90.8 %) [261] | p = 0.182 (2) | p = 0.738 |
| **At least 1 stroke, n (%)** | 7 (13.2 %) [53] | 12 (70.6 %) [17] | 5 (1.9 %) [264] | **p = 0.005** (4) | p = 0.354 (4) |
| **MMSE, mean ± SD** | 29 (27 - 30) [49] | 25 (23 - 29) [17] | *NA* | **p = 0.003** (3) (4) | *NA* |
| **mRS > 1, n (%)** | 1 (2.6 %) [38] | 13 (76.5 %) [17] | *NA* | **p =** NA (5) | *NA* |
| **MMSE ≤ 24, n (%)** | 7 (14.3 %) [49] | 6 (35.3 %) [17] | *NA* | **p = 0.016** (3) (4) | *NA* |
| **Dementia** | 2 (3.8 %) [53] | 2 (11.8 %) [17] | 0 (0 %) [264] | p = 0.276 (4) | p = NA (2) (5) |
| ***CADAMRIT data*** | | | | | |
| **Total WMH score, median (Q1-Q3)** | 1.5 (0 - 3) [40] | 9 (8 - 10) [17] | 2 (1.75 - 4) [24] | **p = $3x10^{-12}$** (4) | p = 0.497 (4) |
| **At least one lacune, n (%)** | 12 (30.8 %) [39] | 14 (82.4 %) [17] | 8 (32 %) [25] | **p = 0.008** (4) | p = 0.447 (4) |
| **At least one microbleed, n (%)** | 2 (5.6 %) [36] | 12 (70.6 %) [17] | 5 (20.8 %) [24] | **p = 0.008** (4) | p = 0.125 (4) |
| **Deep or superficial atrophy, n (%)** | 13 (32.5 %) [40] | 15 (88.2 %) [17] | 12 (48 %) [25] | **p = 0.005** (4) | p = 0.498 (4) |

*Equivalent of Table1 with some comparison regressions on outcomes ((4) and (5)) adjusted differently. For each variable, the number of individuals with available data is reported in square brackets. P-values correspond to the Wald test associated with each variable, adjusting for age (at imaging for CADA-MRIT variables and at inclusion otherwise) and sex, with the following exceptions: (1) not adjusted for age or sex; (2) score test used because the model required for the Wald test did not converge; (3) additionally adjusted for education level; (4) additionally adjusted on hypertension and hypercholesterolemia; (5) the regression additionally adjusted on hypertension and hypercholesterolemia did not converge due to low number of cases. P-values < 0.05 are shown in bold.*
